# Human Phenotype Ontology (HPO) Mapper: Semantic Mapping of Clinical Findings to the Human Phenotype Ontology Using AI-Powered Embeddings and LLM-Based Quality Control

**DOI:** 10.64898/2025.12.20.25342726

**Authors:** Alex Z Kadhim, Zachary Green, Alister Boags, Michael George, Ashley Heinson, Matt Stammers, Christopher M Kipps, R Mark Beattie, Peter N Robinson, James J Ashton, Sarah Ennis

## Abstract

**Background:** Structured phenotypic annotations linked to genetic data can drive diagnostic insight and therapeutic discovery in complex diseases. However, poor research access to the rich clinical data trapped in unstructured clinical records remains a significant barrier to phenotype-genotype integration. Here, we present Human Phenotype Ontology (HPO) Mapper, a scalable AI-assisted tool designed to ingest semantically structured clinical findings paired with anatomical region and accurately map them to HPO terms and associated genes.

**Results:** We applied HPO Mapper to two forms of standardised clinical input extracted from inflammatory bowel disease (IBD) patient records. The first data type consisted of paired ‘clinical findings + anatomical regions’ derived from unstructured clinical reports and the second was standardised ICD-10 code-derived phenotypes. HPO Mapper achieved high semantic alignment and mapping accuracy for both data types (F1 = 0.85 ± 0.05 and 0.84 ± 0.03, respectively). Additionally, HPO Mapper was able to convert 62.3% of previously unusable free-text entries into HPO terms. Utility was demonstrated at cohort scale, whereby resultant HPO sets projected onto gene space recovered well-established IBD drivers including *NOD2*, *IL6*, *STAT3*, *IL10RA*, and *CTLA4*.

**Conclusions:** Our publicly available tool is suitable for converting clinical findings and regions into gene-linked HPO terms for precision medicine. This enables real-time HPO mappings, providing a foundation for scalable AI-driven phenotyping across diseases. More broadly, HPO Mapper provides a generalisable infrastructure for unlocking the latent value of clinical narrative data and bridging the gap between clinical records and genomic diagnostic discovery for targeted therapies.

**VISUAL ABSTRACT:** 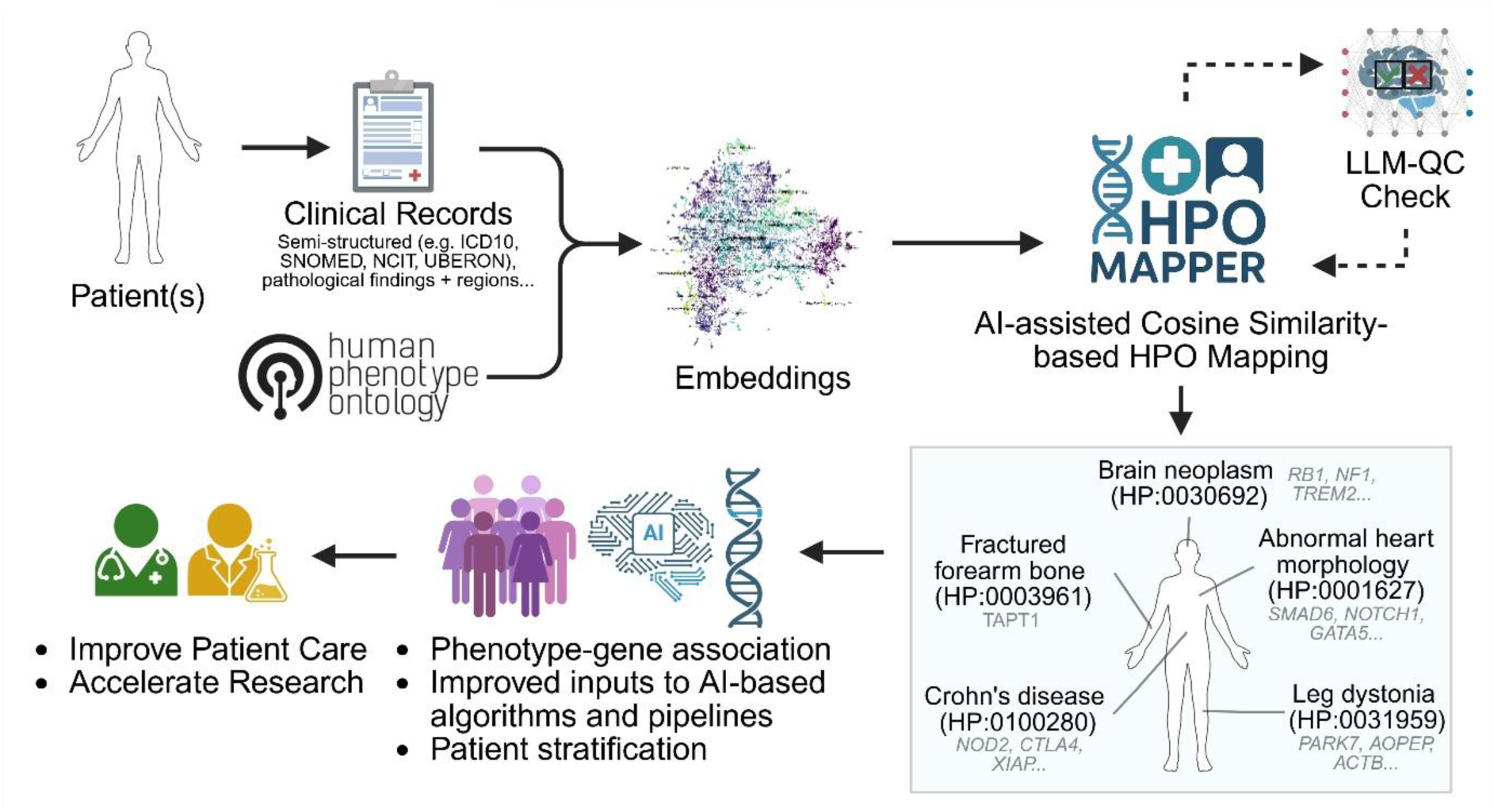

## BACKGROUND

Phenotypic standardisation is a critical prerequisite for effective integration of clinical observations with genomic datasets, especially in complex, multifactorial conditions[1]. Such phenotypic descriptors underpin diagnosis and discovery across rare Mendelian disorders and medical disciplines including neurology, cardiomyopathies, inborn errors of immunity, and inflammatory bowel disease (IBD)[2–6]. Polygenic architecture, modifier effects, and gene-environment interactions generate substantial phenotypic heterogeneity that obscures genotype-phenotype relationships and can limit the diagnostic yield of genomic testing[7,8]. Accurate phenotypic labelling may mitigate this complexity by enabling harmonised cohort aggregation, with phenotype-driven analyses effort such as Exomiser and LIkelihood Ratio Interpretation of Clinical AbnormaLities (LIRICAL) significantly improving variant interpretation and achieving higher diagnostic yield in national-scale programmes[5,9].

The Human Phenotype Ontology (HPO) is a curated, hierarchical, computable vocabulary that standardises the description of human phenotypic abnormalities and links them to diseases, genes, and related ontologies[10]. As a critical resource, HPO enables clinical data to be encoded as machine-readable phenotypes for semantic-similarity analysis, powering phenotype-driven variant and disease prioritisation and enabling interoperable case-level data exchange via the ISO-adopted Global Alliance for Genomics and Health (GA4GH) Phenopacket standard[11,12]. Although HPO provides a structured vocabulary to describe human disease phenotypes[10], clinical data remain predominantly stored in unstructured text formats, such as pathology reports and discharge summaries. Manual structuring of such records is time-consuming, inconsistent, and unscalable. Furthermore, direct mapping approaches often fail to capture the nuanced, context-dependent nature of clinical language[13].

Despite progress in clinical natural language processing (NLP), most existing tools rely on lexicon matching or rule-based pipelines that struggle with semantic matching and the compositional phrases that clinicians use to qualify findings by region, severity or context[14]. While traditional ontology-based approaches, such as the NCBO BioPortal Annotator, provide standardisation, they still tend to favour literal matches and can default to broad parent terms, ultimately reducing specificity in downstream analyses[15]. Recent advances in HPO text mining include dictionary-based systems such as FastHPOCR[16] and alignment-based approaches such as Fenominal[17], which reduce dependence on literal string matching. More recent embedding-based methods that use large language models (LLMs), a type of neural network specialised for text, to capture semantic rather than purely lexical similarity have shown promise in accurate extraction and standardisation[18–20].

By embedding terms and their synonyms into a continuous space where semantic proximity reflects biological relatedness, medical nuance can be preserved[21,22]. LLMs can further arbitrate between close candidates using context, providing a principled route to higher-precision mappings from noisy clinical text into HPO[23,24]. Unfortunately, such models often require higher computational resources, which can be constrained in medical environments, which also have specific privacy requirements[23,25,26]. This motivates designs that precompute ontology embeddings once, apply fast, nearest-neighbour searching for candidate generation, and utilise lightweight, locally deployable models.

Here we introduce HPO Mapper, a modular framework that combines embedding-based candidate generation with targeted LLM mediation. We present three protocols that incrementally incorporate LLM support, from embedding-only selection to LLM quality control, to LLM-assisted selection with quality control. We first show that HPO embeddings preserve biological structure, then evaluate mapping performance on an IBD-patient derived clinical dataset. Finally, we demonstrate downstream value by linking mapped HPO terms to genes and pathways across 32,041 clinical records, recovering known IBD mechanisms and highlighting ontology gaps that merit curation. Together, these contributions provide an accurate, scalable and privacy-preserving route to phenotype standardisation for integrative genomics.

## IMPLEMENTATION

### Cohort and NHS electronic health record

We obtained clinical data from a total of 1,837 research-consented patients diagnosed with inflammatory bowel disease recruited through the University of Southampton paediatric and adult clinics as part of the ethically approved Southampton Genetics of IBD Study (REC 09/H0504/125; Table 1). All consenting participants meeting these criteria were included without further sampling or selection. Clinical records were redacted using *Pteredactyl*, a publicly available clinical free-text redaction tool designed to remove personally identifiable information including dates, patient and clinician names [24,27].

**Table 1:** Summary characteristics of studied cohort.

| Demographic variable | Total (percentage, %) or median, decimalised years (range) |
| --- | --- |
| Total participants | 1837 |
| Male | 950 (51.7%) |
| Female | 887 (48.3%) |
| Crohn's disease | 1127 (61.4%) |
| Ulcerative colitis | 667 (36.3%) |
| IBDU | 25 (1.4%) |
| Unknown/Other | 18 (1.0%) |
| Diagnosed <17 years | 773 (42.1%) |
| Diagnosed ≥17 years | 987 (53.7%) |
| Missing age at diagnosis | 77 (4.2%) |
| Age at IBD diagnosis (years), median (range) | 19.6 (0.1–88.8) |
| Total follow-up duration | 18.2 (0.0–29.4) |

### Data sources

To assess initial HPO Mapper performance, we evaluated two distinct real-world data types as input datasets obtained from a longitudinally assessed cohort of IBD patients. The first included paired ‘clinical findings + anatomical regions’ extracted from histology and imaging reports processed in a standardised manner, as previously described[24]. A total of 32,041 unstructured reports were available, comprising histological procedures (n = 7,181; 22.41%) and imaging procedures (n = 24,860; 77.59%). Imaging covered 13 modalities, including ultrasound (21.02%), magnetic resonance imaging (17.25%), X-ray (14.58%), computed tomography (12.52%), and legacy uncoded reports (17.25%), among others.

The second data input was derived from the set of standardised ICD10 codes (Table 2) extracted from longitudinal hospital discharge summaries. This yielded 50,489 timestamped ICD10 codes across the cohort of 1,837 patients. Z999 ICD10 codes (n = 8,823), which represent free-text ICD10 entries, are typically selected when the correct diagnostic code cannot be readily identified. These codes often contain spelling errors or ambiguous terminology and occur more frequently in contexts of high clinical workload. The study remains active with a median follow-up duration of 11.62 years.

**Table 2:** International Statistical Classification of Diseases and Related Health Problems 10th Revision (ICD10) Chapters and Titles.

| Prefix | Chapter Name | Examples |
| --- | --- | --- |
| **A/B** | Certain infectious & parasitic diseases | Infectious gastroenteritis and colitis, unspecified (A09.0)\|Respiratory tuberculosis, confirmed (A15.0)\|Human immunodeficiency virus [HIV] disease (B20) |
| **C** | Neoplasms | Malignant neoplasm of colon, unspecified (C18.9)\|Malignant neoplasm of breast, unspecified (C50.9)\|Malignant neoplasm of bronchus and lung, unspecified (C34.9) |
| **D** | Neoplasms; Blood & immune disorders | Lymphoid leukaemia, unspecified (C91.9)\|Iron deficiency anaemia, unspecified (D50.9)\|Common variable immunodeficiency, unspecified (D83.9) |
| **E** | Endocrine, nutritional & metabolic | Type 2 diabetes mellitus without complications (E11.9)\|Hypothyroidism, unspecified (E03.9)\|Hyperlipidaemia, unspecified (E78.5) |
| **F** | Mental & behavioural disorders | Depressive episode, unspecified (F32.9)\|Schizophrenia, unspecified (F20.9)\|Alcohol dependence syndrome (F10.2) |
| **G** | Diseases of the nervous system | Epilepsy, unspecified (G40.9)\|Migraine, unspecified (G43.9)\|Multiple sclerosis (G35) |
| **H** | Eye & adnexa; Ear & mastoid | Unspecified conjunctivitis (H10.9)\|Senile cataract, unspecified (H25.9)\|Otitis externa, unspecified (H60.9) |
| **I** | Diseases of the circulatory system | Acute myocardial infarction, unspecified (I21.9)\|Essential (primary) hypertension (I10)\|Cerebral infarction, unspecified (I63.9) |
| **J** | Diseases of the respiratory system | Asthma, unspecified (J45.9)\|Pneumonia, unspecified organism (J18.9)\|Chronic obstructive pulmonary disease, unspecified (J44.9) |
| **K** | Diseases of the digestive system | Crohn's disease, unspecified (K50.9)\|Ulcerative colitis, unspecified (K51.9)\|Gastro-oesophageal reflux disease without oesophagitis (K21.9) |
| **L** | Skin & subcutaneous tissue | Atopic dermatitis, unspecified (L20.9)\|Psoriasis, unspecified (L40.9)\|Cellulitis, unspecified (L03.9) |
| **M** | Musculoskeletal & connective tissue | Osteoarthritis of hip, unspecified (M16.9)\|Rheumatoid arthritis, unspecified (M06.9)\|Low back pain (M54.5) |
| **N** | Genitourinary system | Chronic kidney disease, unspecified (N18.9)\|Urinary tract infection, site not specified (N39.0)\|Endometriosis, unspecified (N80.9) |
| **O** | Pregnancy, childbirth & puerperium | Gestational hypertension without significant proteinuria (O13)\|Gestational diabetes mellitus, diet controlled (O24.4)\|Single spontaneous delivery (O80) |
| **P** | Perinatal conditions | Extremely low birth weight newborn (P07.0)\|Respiratory distress syndrome of newborn (P22.0)\|Neonatal jaundice, unspecified (P59.9) |
| **Q** | Congenital malformations & chromosomal abnormalities | Ventricular septal defect (Q21.0)\|Down syndrome, unspecified (Q90.9)\|Spina bifida, unspecified (Q05.9) |
| **R** | Symptoms, signs & abnormal findings | Chest pain, unspecified (R07.4)\|Abdominal pain, unspecified (R10.9)\|Headache (R51) |
| **S/T** | Injury, poisoning & consequences of external causes | Fracture of forearm, unspecified (S52.9)\|Poisoning by systemic antibiotics (T36.0)\|Injury, unspecified (T14.9) |
| **U** | Codes for special purposes | COVID-19, virus identified (U07.1)\|COVID-19, virus not identified (U07.2)\|Post COVID-19 condition, unspecified (U09.9) |
| **V/W/X/Y** | External causes of morbidity & mortality | Pedestrian injured in collision with car, pick-up truck or van (V03.1)\|Unspecified fall (W19)\|Intentional self-harm by hanging, strangulation and suffocation (X70) |
| **Z** | Factors influencing health status & contact with health services | Encounter for chemotherapy (Z51.1)\|Screening for neoplasm of intestinal tract (Z12.1)\|Need for immunisation (Z23) |

### Semantic embedding of HPO and branch similarity

For creation of the reference database, all HPO v2025-09-01 terms (19,903) and synonyms (24,161) were converted into embeddings using *nomic-embed-text:v1.5[28]*. Embeddings are numerical representations of text that encode semantic meaning, enabling comparison of concepts based on their contextual similarity rather than exact lexical overlap[29]. This corresponded to a mean of 1.21 synonyms per term and a median of 1. Notably, 8,872 terms (44.6%) had no synonyms. For visualisation purposes, HPO terms were restricted to descendants of HP:0000118 (Phenotypic abnormality; Supplementary Figure 1B) by restricting terms to descendants of HP:0000118. A 2D and 3D interactive plot of embeddings can be viewed at: https://huggingface.co/spaces/UoS-HGIG/HPOmapper.

### HPO embedding validation

To test whether converted embeddings reflect biology, we computed pairwise distances in the original embedding space for a stratified set of term pairs, labelling each as within-category (intra) or across-category (inter). For example, ‘Hematochezia’ (HP:0012573) and ‘Rectal bleeding’ (HP:0002575) are intra and should be closer than ‘Perianal fistula’ (HP:0100518) and ‘Uveitis’ (HP:0000554), which are inter. Embeddings were L2-normalised and cosine distance (*1 − cosine similarity)* was used throughout[30]. A two-dimensional projection was generated using t-SNE after a principal component analysis (PCA) pre-step, points were coloured by top-level branch. The distributions of embedding distances between terms were visualised using violin plots, and a branch comparison heatmap was assembled by taking the mean cosine distance for each category pair (diagonal = within-category).

As some HPO terms occur in multiple branches, we used inter-category distances as a null model to test whether intra-category embedding distances were smaller than expected by chance. For each HPO term we measured the cosine similarity distance between it and every other HPO term using a one-sided Mann-Whitney U test. This was performed to confirm that the HPO-derived semantic embeddings faithfully recreate the ontological hierarchy. We tested the hypothesis that a given category pair has significantly smaller distances (i.e. greater semantic similarity) than across categories. P values were subsequently adjusted for multiple testing using Benjamini-Hochberg false discovery rate (FDR) correction. All steps were executed in Python using NumPy, Pandas, and scikit-learn/Matplotlib.

### Evaluation and metrics

To assess the performance of our HPO mapper protocols, we manually curated reference gold-standard corpus containing 4,112 statements formed from 3,290 (training) and 822 (testing) finding and region pairs. A randomly selected set of findings and regions was run through HPO Mapper Protocol 1 (P1) and manually reviewed by a clinical gastroenterologist (ZG). The manually curated output was then randomly divided, using 80% for training and procedure tuning and holding out 20% strictly for final evaluation. Model output for each of the 822 test input statements was compared against a single gold standard HPO ID term that had been manually curated by a clinical expert. Predictions were scored as follows: true positive when the predicted HPO ID exactly matched the gold standard ID, counting HPO synonyms as the same ID; false negative when a non-empty prediction was returned with a non-matching ID; false negative when no prediction was returned. By convention a wrong prediction contributes one FP and one FN, while an unmapped item contributes one FN. We report macro-averaged precision, recall, and F1-score across all test statements, where:

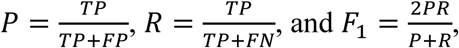

with 𝑇𝑃, 𝐹𝑃, and 𝐹𝑁denoting true positives, false positives, and false negatives, respectively.

### HPO Mapper and protocols evaluated

All clinical queries (findings and regions) were embedded using the *nomic-embed-text:v1.5* model via the Ollama python package (v0.6.0)[24,28]. Across all protocols, multi-region statements were split into separate “{finding} and {region}” items before mapping. If a region was absent only the finding was mapped. Generated embeddings were then compared against precomputed embeddings for canonical and synonym HPO term[24].

Cosine similarity was used to identify the most similar HPO concept. As previously determined by threshold optimisation[24], only matches with cosine similarity > 0.76 were retained.

Three different protocols were evaluated for optimal mapping of input statements to HPO terms. These trials were performed on the more complex data type where input statements were the paired ‘clinical findings + anatomical regions’. The optimal protocol was then deployed on a large dataset of standardised ICD-10 codes. Protocol 1 (P1) used candidate retrieval with string and embedding similarity against an index that included HPO terms and synonyms. Exact or near exact matches (cosine similarity > 0.76) to preferred HPO terms or synonyms terms were prioritised. Retrieval returned a ranked list of candidates with cosine similarity scores. In cases with ties, the higher-level parent term was selected, when appropriate. No LLM was used in this protocol.

Protocol 2 (P2) added an LLM based quality control step on top of P1 retrieval. Table 3 lists all LLMs compared in this study. A fine-tuned LLM received each input statement from P1 and their definitions, then validated the leading choice or proposed a correction. The final fine-tuning set comprised 4,112 ‘finding and region’ pairs that were first mapped by P1 and then manually corrected by a clinical expert (ZG). To assess model choice, the LLM-QC checker component of P2 was instantiated in turn with the following LLM backends under identical prompts and decision policy: gpt oss, Llama 3.3, Gemma 3, DeepSeek R1, Mistral, and Llama 3.1 8B. The core prompt was as follows:

**Table 3:** LLMs utilised in study.

| LLM | Size (GB) | Variation |
| --- | --- | --- |
| **llama3.1** | 16 | llama3.1:8b-instruct-fp16 |
| **llama3.3** | 141 | llama3.1:70b-instruct-fp16 |
| **deepseek-r1** | 43 | deepseek-r1:70b |
| **mistral-large** | 130 | mistral-large:123b-instruct-2407-q8_0 |
| **gpt-oss** | 65 | gpt-oss:120b |
| **gemma3** | 17 | gemma3:27b |

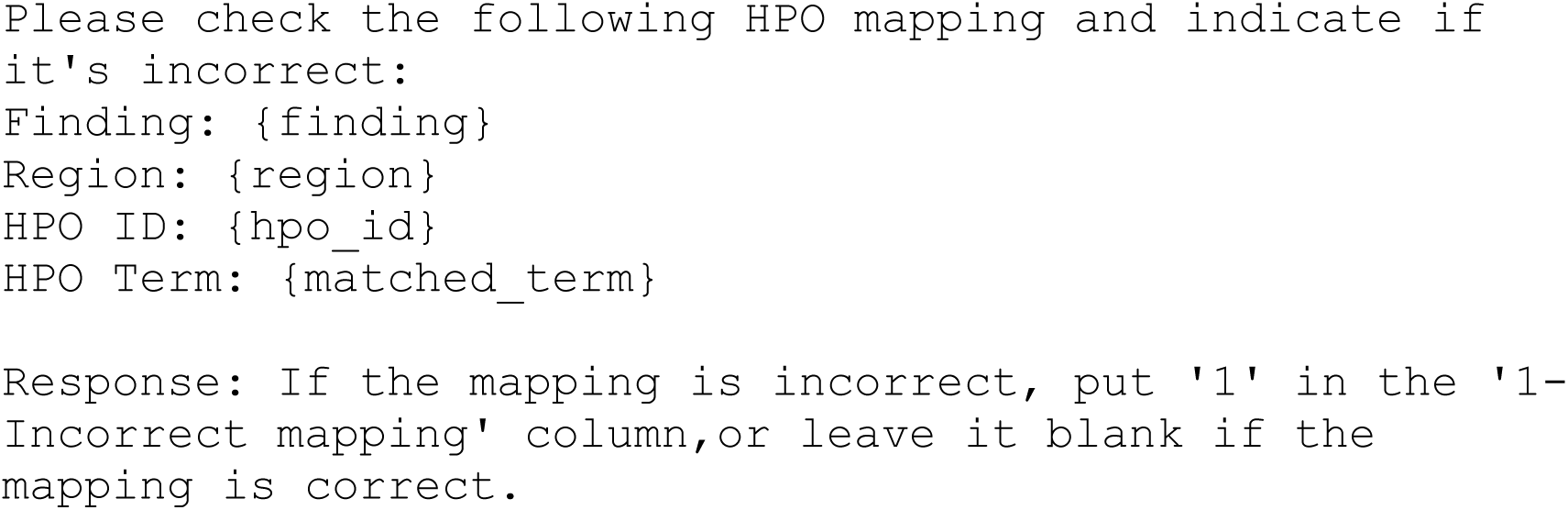

Protocol 3 (P3; LLM assisted term matching plus LLM QC) first used P1 to produce the top 10 candidate matches for each input statement. A base LLM was then presented with the statement, 10 candidates, and their definitions, and asked to select two terms it judged closest. Each selected term was subsequently passed through the P2 quality control check component. We evaluated the same set of LLMs assisted term matching as described in P2. If both selected terms were rejected by QC the item was marked unmapped. If one or both passed QC the passing term was retained, with the first QC approved selection used for primary analyses. The core prompt used was as follows:

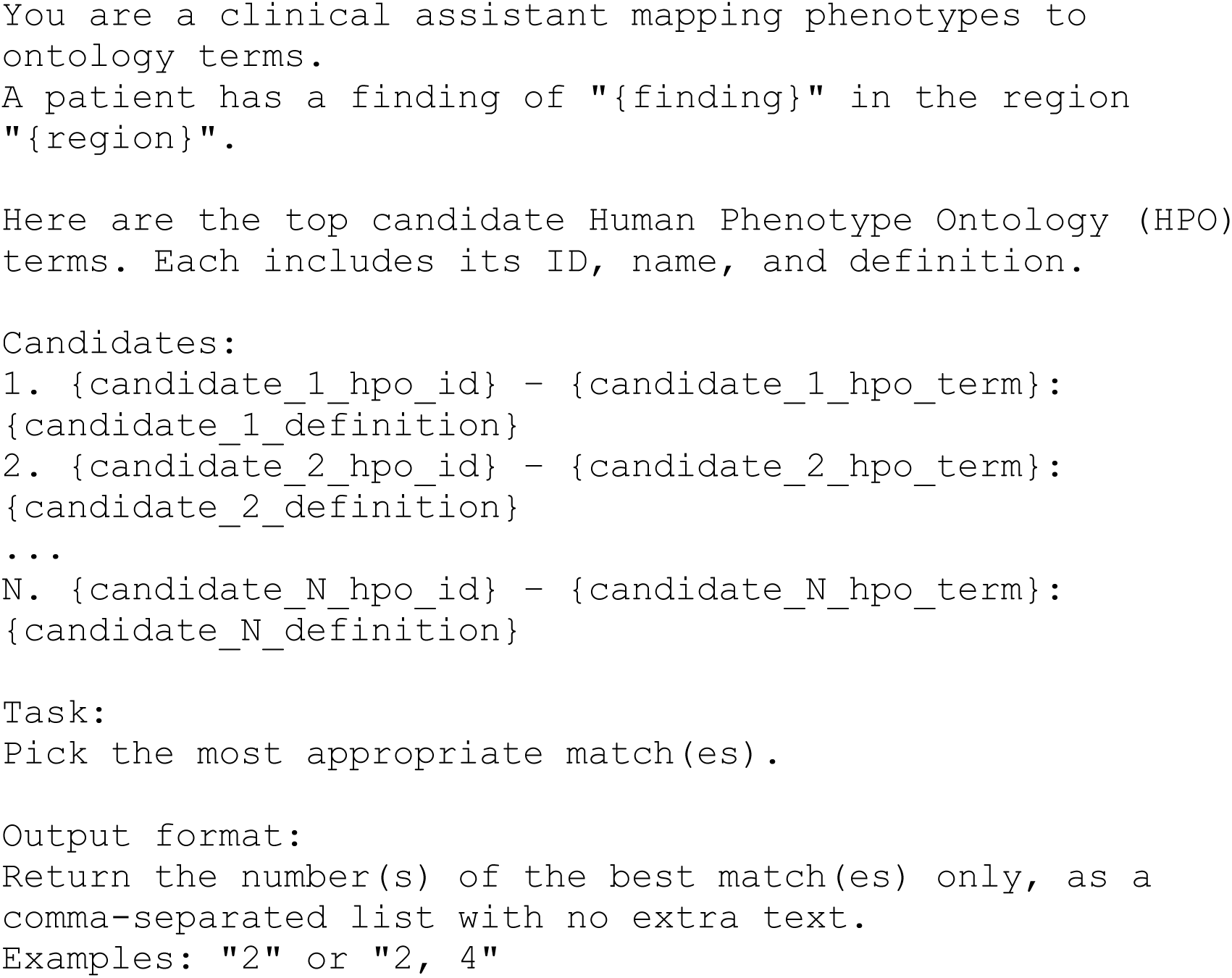

### Benchmarking against current HPO mapping tools

We called the BioPortal Annotator REST API from Python using the requests library as an external ontology tagger annotator baseline tool. Each input statement was constructed from finding plus region in the form “{finding} in {region},” or from the finding alone when the region was absent, after lowercasing, Unicode normalisation, and light punctuation cleanup. Requests targeted the Annotator endpoint with ontologies set to HP, format set to JSON, and default parameters except longest_only set to true. API credentials were supplied via an environment variable, and submissions were sent in small batches with retry logic and exponential backoff on transient errors or rate limits. The JSON response was parsed to extract HPO identifiers (containing “HP:” prefix), preferred labels, offsets, and annotator scores. When multiple HP concepts were returned for a statement, we retained the highest scoring concept; in the case of tied scores, we selected the concept with greater specificity based on ontology depth when available, otherwise the first returned. For each processed statement we recorded the chosen HP identifier, label, and score. Statements with no HP matches were marked unmapped.

A set of UMLS mapped ICD10 codes and patient-derived histology/radiology terms were mapped to HPO using fenominal, FastHPOCR, and HPO-RAG [16,17,31]. ICD10 inputs consisted of code labels extracted from the UMLS-derived ICD10/ICD10-CM-to-HPO shared-CUI mapping table (n = 3,601). Patient inputs consisted of individual structured entries, obtained as previously described[24], containing a finding, anatomical regions, and qualifiers. These were combined into a single text string per entry. Fenominal and FastHPOCR were run on one plain-text file per input. Fenominal was executed using the Java command-line parser with hp.json, and excluded matches were removed before evaluation.

FastHPOCR was executed with a pre-built HPO index and the HPOAnnotator, with outputs serialised to tab-separated files. HPO-RAG was run from the RAG-HPO github page (https://github.com/PoseyPod/RAG-HPO) using csv inputs containing patient_id and clinical_note, with a mapping table used to link predictions back to the original input identifiers. HPO-RAG was connected to a local Ollama-hosted language model (Llama 3.1-70b) on an AMD Instinct^TM^ MI300X GPU, using an offline tokeniser patch to avoid internet-dependent tiktoken downloads and an increased token limit for large batched runs. All tool outputs were normalised into a shared long-format table containing input key, tool, HPO ID, HPO label, and matched text. Predictions were compared with the gold-standard HPO mappings using precision, recall, F1 score, per-input macro metrics, and standard deviations across inputs as previously described[24]. Runtime was assessed on stratified random samples of ICD10 and patient-derived terms grouped by text length.

### Gene association mapping

Each mapped HPO term was linked to its associated genes using the curated HPO phenotype to gene table (phenotype_to_genes.txt) obtained from hpo.jax.org[1]. We parsed the table to retrieve gene symbols and corresponding identifiers for each HPO ID observed in our data. Gene symbols were normalised to current HGNC preferred symbols where possible, and deprecated aliases present in the file were retained as secondary labels for traceability.

For each patient we aggregated the union of genes across all mapped HPO terms, removed duplicates, and recorded both per statement gene links and per patient summaries. The primary output was a long format CSV with columns that included patient identifier, source statement, mapped HPO ID and label, and gene symbol. From this table we derived patient level metrics that included the number of unique mapped HPO terms, the number of unique genes associated with those terms, and the frequency with which each gene appeared across a patient’s statements. If an HPO term had no listed gene associations in the table it contributed no genes but remained in the HPO summary.

### Genes to pathway associations

Patient level gene sets derived from the HPO mappings were analysed with Enrichr through its programmatic interface, using the default human gene background. We queried Enrichr disease and pathway libraries[32], including disease ontologies and pathway collections such as Disease Ontology, DisGeNET, Reactome, and KEGG, to identify enriched terms for each patient specific gene set[33–35]. Enrichment used Fisher’s exact test with Enrichr’s combined score, and p values were adjusted across all tested terms per library using the Benjamini-Hochberg procedure. Unless noted otherwise, results were considered significant at FDR < 0.05 with a minimum overlap of two genes. For each patient and library, we exported a ranked table with term name, source library, overlap, odds ratio, p value, adjusted p value, and combined score. We then used Enrich-KG[36] to generate a knowledge graph that linked patients to their HPO terms, HPO terms to genes, and genes to enriched diseases and pathways. Edge weights captured the strength of evidence, for example overlap counts for HPO to gene links and negative log_10_ adjusted p values for enrichment links. The graph was exported in standard formats for downstream visualisation and auditing.

### HPO to gene ranking and causal gene list generation

Histology and imaging reports from three patients with confirmed pathological variants in genes *CTLA4*, *XIAP*, and *FOXP3* respectively were extracted and standardised using a previously published methodology [24]. Outputs, including additional ICD10 coded comorbidities, were converted to HPO terms using HPO Mapper P2. To rank potentially causal genes, we calculated an HPO-based phenotype-genotype similarity score (Patient Gene HPO Overlap) for each gene-patient pair using the following formula:

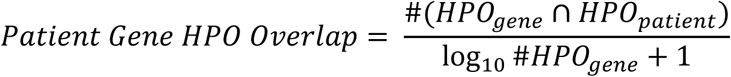

The denominator allow correction for genes that contain higher numbers of phenotypic annotations in HPO. For gene prioritisation within each patent, gene lists were sorted by score and min-max scaled (n = 5,252 genes currently annotated by HPO). Similarly, min-max scaling was applied to each gene across the entire cohort of IBD patients (n = 1,573) to visualise location and distribution within the cohort.

## RESULTS

### Embeddings of HPO are biologically relevant and can be used for downstream matching

To assess the performance of HPO mapper protocols on real-world data, we tested against a manually curated dataset (n = 822) of finding-region pairs obtained from imaging and histology reports from IBD patients. In total, we evaluated three HPO mapper protocols against the NCBO BioPortal Annotator tool using cosine-similarity selection. Protocol 1 performs cosine similarity matching on input to HPO terms; Protocol 2 adds an LLM-based quality control step that accepts or rejects the chosen term; Protocol 3 allows an LLM to assist with the initial term selection before applying the same quality control (Figure 1A).

**Figure 1:**
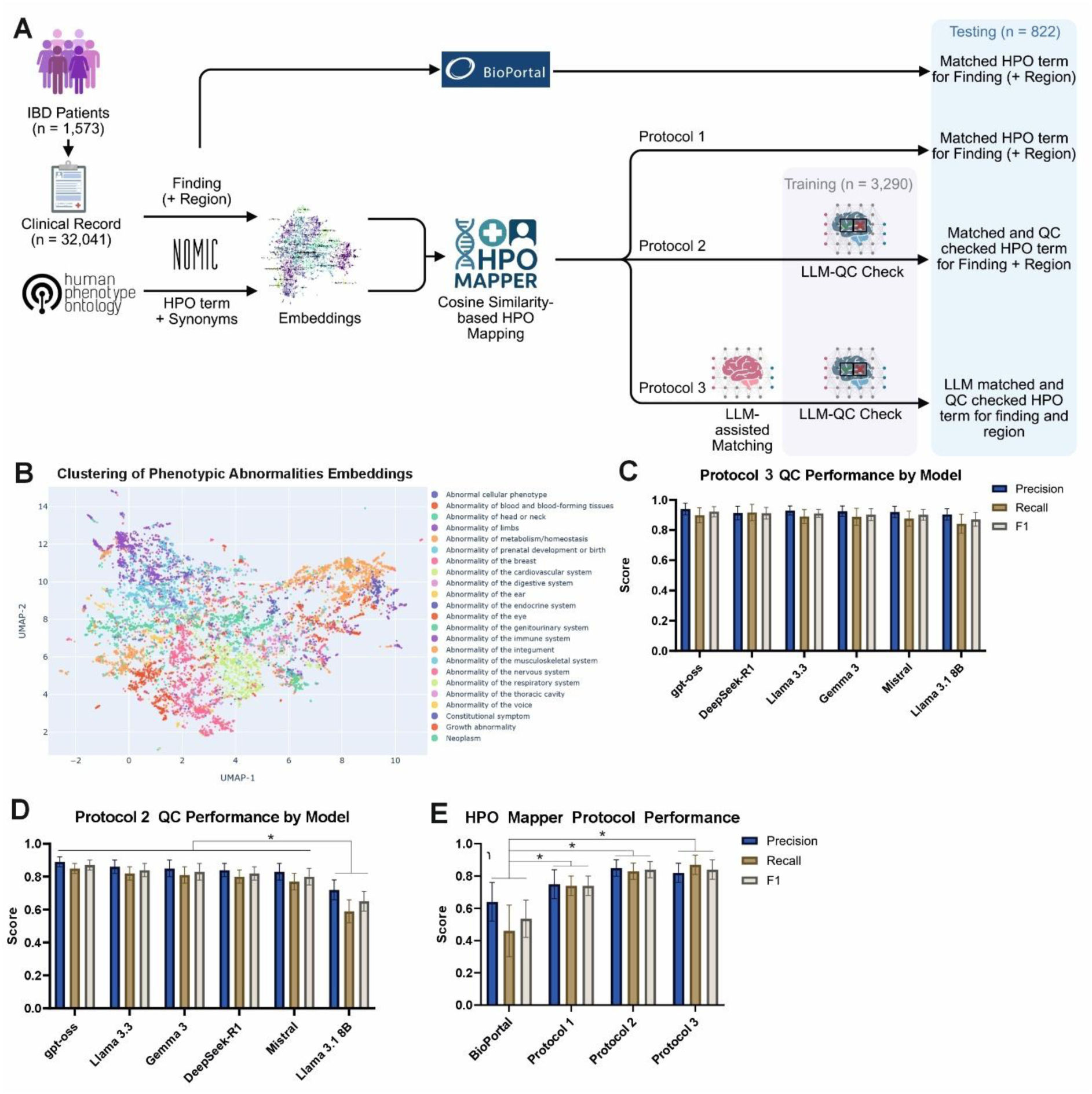
HPO embeddings cluster into distinct biologically meaningful groups and allow matching using large language models. A) Schematic detailing input and outputs of HPO Mapper Protocol 1 (P1), 2 (P2), and 3 (P3). B) t-SNE plot of HPO and synonym embeddings showing HPO terms under “Phenotypic abnormality” (HP:0000118) branch converted by Nomic. C) Comparison of LLMs in P2-assisted QC checking of HPO matches. D) Comparison of LLMs in P3-assisted selection of HPO matches. E) Comparison of HPO mapper performance between Bioportal, Protocol 1 (P1), Protocol 2 (P2), and Protocol 3 (P3). All data are shown as mean ± SEM.

Prior to testing HPO mapper, we assessed the usability of HPO embeddings. A PCA projection was generated under the “Phenotypic Abnormality” branch (HP:0000118) and clusters forming coherent and biologically relevant organ-system neighbourhoods were observed (Figure 1B). A magnified view shows contiguous areas for abnormalities of the digestive, respiratory, and cardiovascular systems, with clear internal structure (Supplementary Figure 2A). As expected, distances between HPO terms with a shared parent category (intra-term distances) were significantly smaller (p value < 0.001), whereas values between HPO terms from different categories (inter) being higher (Supplementary Figure 2B). Inter-branch relationships revealed biologically meaningful proximities among HPO term embeddings, with closely related parent terms showing strong similarity while unrelated pairs (e.g., immune vs. ear abnormalities) did not (Supplementary Figure 2C). These results indicate that the embedding space preserves semantic relatedness and concentrates closely related phenotypes when projected back to HPO and is therefore suitable for matching.

### HPO mapping can be improved with LLM-assisted mediation

To assess the effect of LLM choice as a quality control agent (Protocol 2), we fine-tuned six different models on a training set of 3,290 validated finding-region HPO matches. Models were presented with an input (finding and/or region) and a single candidate HPO match to flag as incorrect. All tested LLMs delivered high precision, recall, and F1 (> 0.8), indicating stable behaviour of the quality control step (Figure 1C). Relative to the embedding-only approach, introducing quality control raised precision while maintaining strong recall, suggesting that most gains came from removing false positives rather than discarding correct matches.

We next determined whether LLMs could be used to select the best match(es) when presented with the top 10 HPO matches and definitions using cosine similarity (Protocol 3). Performance of the LLM-assisted HPO matching Protocol 3 remained high across larger-parameter models, and gpt-oss had the highest overall performance (F1 = 0.87 ± 0.03; Figure 1D). Llama-3.1-8B performed significantly worse compared to other models across metrics (F1 = 0.65 ± 0.06, p value < 0.05). This suggests that larger models may perform better, when used for LLM-assisted HPO matching.

When results were aggregated by protocol using gpt-oss for Protocols 2 and 3, both protocols matched or exceeded the BioPortal baseline across all metrics (Figure 1E). Protocol 2 and Protocol 3 performed significantly better than BioPortal for both recall and F1 score (p = 0.02 and p = 0.01), while differences in precision were not statistically significant. Protocols 2 and 3 showed highly similar overall performance, with no significant difference in F1 score (p = 0.67), indicating that both methods deliver comparably strong accuracy.

Overall, Protocol 2 provided the best balance of precision, recall, and computational requirements, showing that targeted LLM checks and LLM-assisted selection improve HPO mapping quality without loss of recall, while also providing a computationally efficient solution compared to Protocol 3.

### ICD10 coded patient diagnoses can be reliably converted to HPO using HPO mapper

To evaluate ICD10 to HPO conversion at scale, we analysed ICD10-coded diagnoses derived from the clinical records of 1,837 IBD patients, comprising 50,489 codes, and applied HPO Mapper Protocol 2 to produce HPO terms (Figure 2A). For comparisons between mapping methods, we included a direct conversions that first maps ICD10 codes to SNOMED CT and then to HPO, as well as the NCBO BioPortal Annotator. The distribution of codes in this IBD cohort was enriched for gastrointestinal and symptom codes, with Crohn’s disease and ulcerative colitis among the most frequent entries, alongside abdominal pain, non-infective gastroenteritis, iron deficiency anaemia, and treatment or history codes (Figure S3A). This pattern underlines the importance of capturing both disease labels and symptomatology when converting ICD10 to HPO.

**Figure 2:**
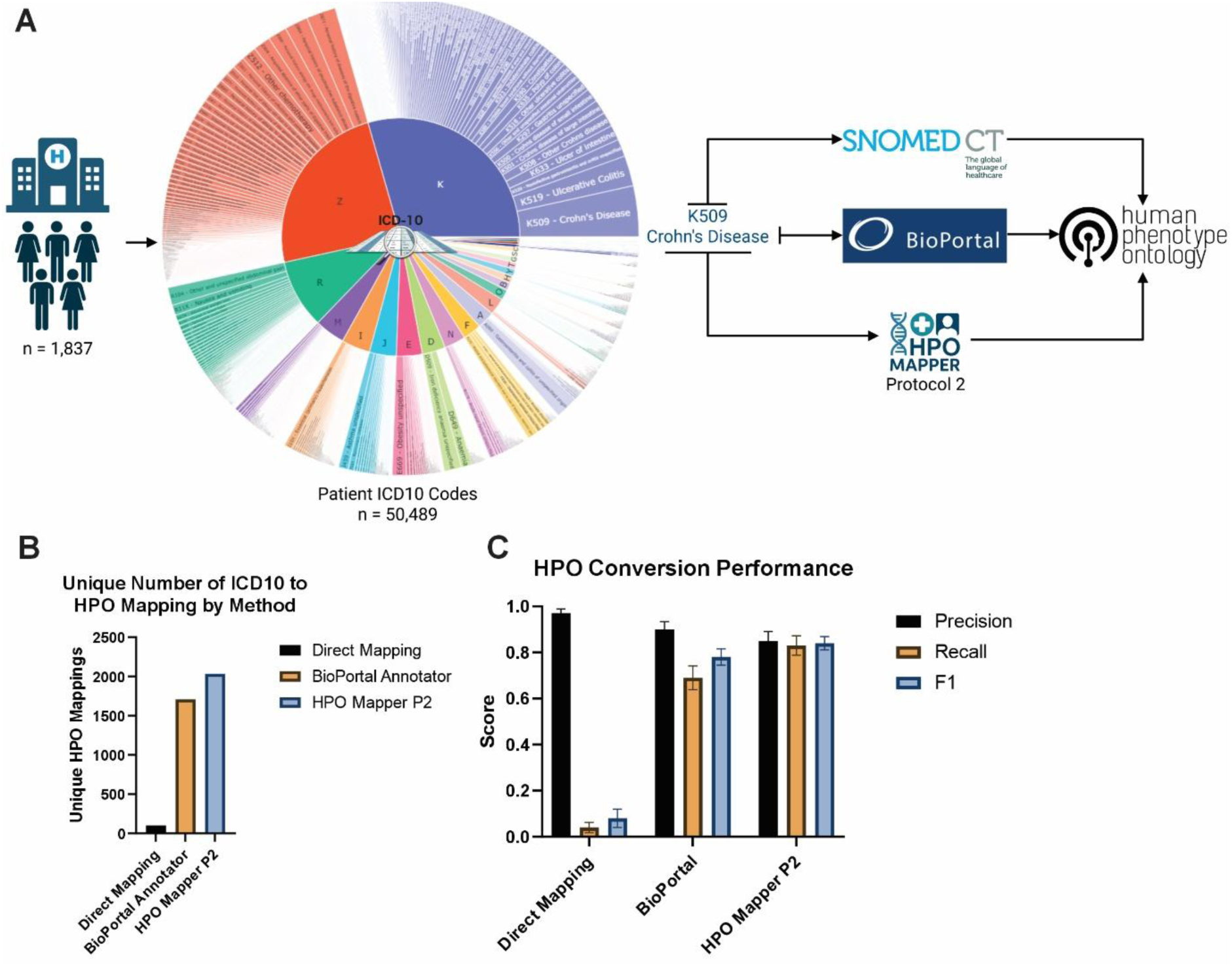
HPO Mapper Protocol 2 improves ICD10 conversion to HPO terms. A) Schematic of data processing pipeline using IBD patient (n = 1,837) obtained ICD10 codes (n = 50,489) through SNOMED CT (Direct Mapping), NCBO BioPortal Annotator, or HPO Mapper Protocol 2 (P2). B) Unique number of ICD10 codes converted to HPO using direct mapping (ICD10 to SNOMED to HPO), BioPortal Annotator API, or HPO Mapper P2 (n = 3,601). C) Performance of the three described annotation methods measured by macro precision, recall, and F1 score. All data are shown as mean ± SEM.

Across mapping methods, HPO Mapper P2 yielded the highest number of unique HPO terms from ICD10 codes, followed by BioPortal Annotator, and direct mapping (Figure 2B). Similarly, the direct conversion of ICD10 to SNOMEDCT and HPO showed high precision but reduced recall, reflecting its limited catalogue of explicit mappings, while BioPortal improved recall but had a lower F1 score (Figure 2C). HPO Mapper Protocol 2 achieved the best combined precision and recall and therefore the highest F1 (0.84±0.03), indicating that synonym expansion with embeddings and an LLM quality control step captures phenotypic signal missed by rule-based approaches, yielding broader and more clinically meaningful HPO coverage without loss of accuracy (Figure 2C).

We next assessed HPO Mapper P2’s performance to other current tools for HPO term extraction and conversion[16,17,31]. Using the full set of UMLS-derived ICD10/ICD10-CM HPO mappings (n = 3,601), both HPO Mapper P2 and HPO-RAG scored highest with F1 scores of 0.866 and 0.870 respectively (Table 4A). Both Fenominal and FastHPOCR demonstrated lower performance (F1 = 0.762 and 0.705, respectively) but demonstrated an average of 1.83x faster processing speed and lower power consumption per input compared to HPO Mapper 2 or HPO-RAG (Supplementary Figure 3). When converting a subset of standardised finding region pairs from histology and radiology reports (n = 1,118), HPO Mapper P2 had improved performance compared to HPO-RAG, Fenominal, and FastHPOCR (Table 4B) with an average F1 of 0.837 for HPO Mapper P2 compared to a combined average F1 of 0.595 for all other tools. Taken together, this demonstrates that HPO Mapper P2 is comparable to other current HPO conversion tools.

**Table 4:**
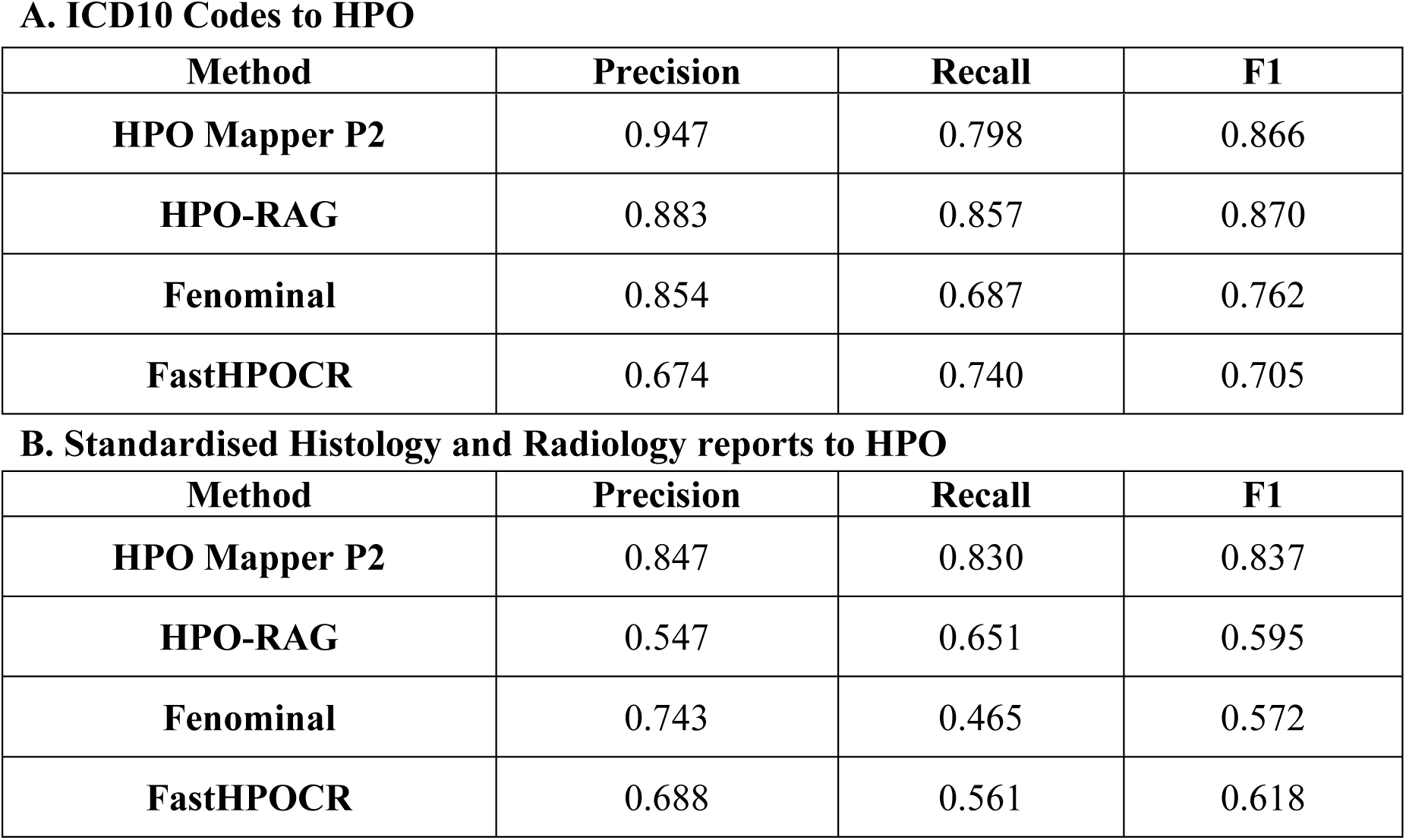
Performance of HPO Mapper P2, HPO-RAG, Fenominal, and FastHPOCR in converting A) a validated subset of ICD10 codes (n = 3,601) and B) standardised histology and radiology report finding and region pairs (n = 1,118), to HPO by micro-average precision, recall, and F1.

**A. ICD10 Codes to HPO**
| Method | Precision | Recall | F1 |
| --- | --- | --- | --- |
| **HPO Mapper P2** | 0.947 | 0.798 | 0.866 |
| **HPO-RAG** | 0.883 | 0.857 | 0.870 |
| **Fenominal** | 0.854 | 0.687 | 0.762 |
| **FastHPOCR** | 0.674 | 0.740 | 0.705 |

**B. Standardised Histology and Radiology reports to HPO**
| **Method** | **Precision** | **Recall** | **F1** |
| --- | --- | --- | --- |
| **HPO Mapper P2** | 0.847 | 0.830 | 0.837 |
| **HPO-RAG** | 0.547 | 0.651 | 0.595 |
| **Fenominal** | 0.743 | 0.465 | 0.572 |
| **FastHPOCR** | 0.688 | 0.561 | 0.618 |

ICD10 to HPO coverage by HPO Mapper P2 was further examined by segmenting analysis into ICD10 chapters to reflect the breadth of clinical content represented in our hospital record set. In both our IBD cohort and the entire ICD10 dictionary, disease-focused chapters showed high mapping rates, with most chapters exceeding 70% coverage (Figure 3A). Neoplasms, endocrine disorders, and digestive system disorders achieved the highest coverage among other ICD10 chapters. As expected, chapters that are administrative or context-oriented rather than phenotypic, such as external causes of morbidity, factors influencing health status, and special purposes, had minimal coverage, with external causes close to zero and special purpose codes unmapped (Figure 3A).

**Figure 3:**
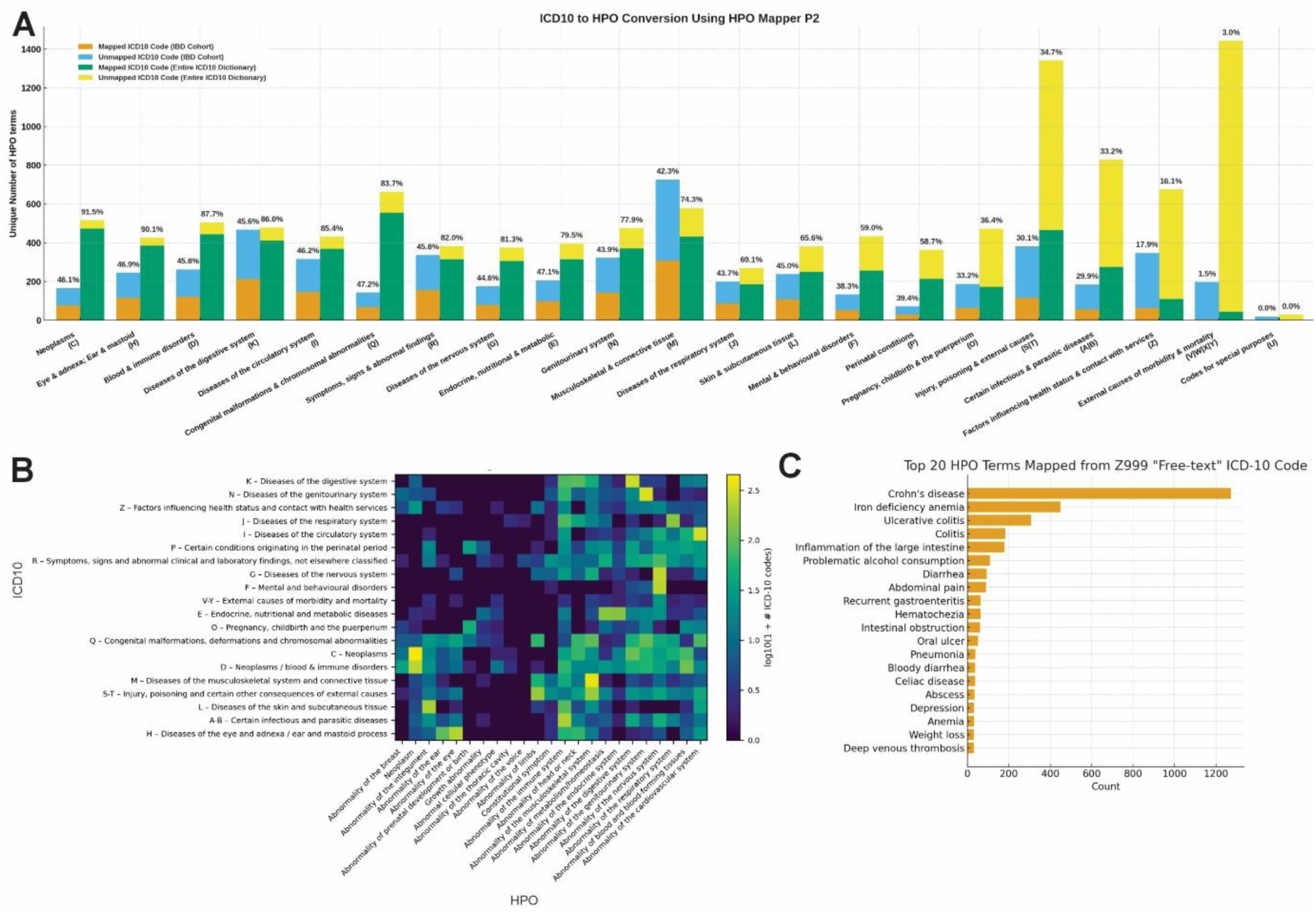
HPO Mapper Protocol 2 can map patient histology and radiology reports to HPO terms. A) Unique number of HPO mapped and unmapped ICD10 codes obtained from IBD Cohort and ICD10 Dictionary grouped by ICD10 prefix using HPO Mapper P2. B) Heatmap of ICD10 to HPO conversion showing distribution of output HPO term by parent class. C) Top 20 HPO terms derived from free-text ICD10 (Z999) codes.

Among ICD10 chapter prefix groups to HPO parent terms, we observed a high degree of expected outputs such as Diseases of the digestive system (K) to Abnormality of the digestive system (HP:0025031; 58.59% of all K ICD10 prefix terms) and Neoplasm (C) with Neoplasm (HP:0002664; 84.60% of all C ICD10 prefix terms; Figure 3B). Lastly, HPO Mapper P2 was able to convert Z999 free-text ICD10 codes (n = 8,823), typically selected when the correct diagnostic code cannot be readily identified due to ambiguous terminology, high clinical workload, or limited coding experience, into usable HPO terms. HPO Mapper P2 yielded additional mappings for Crohn’s disease (HP:0100280; 1272 counts), Iron deficiency anemia (HP:0001891; 450 counts), and Ulcerative colitis (HP:0100279; 308 counts), among other Z999 free-text entries (Figure 3C), leading to 5,498 additional HPO terms (62.3% of all Z999 codes). These findings demonstrate the usability of HPO Mapper in converting structured and otherwise unusable unstructured free-text input into usable HPO terms for downstream analysis.

### HPO term to gene mappings reveal biologically meaningful signals

To determine if biological insight could be extracted from linked HPO and gene associations, we processed all clinical records from IBD patients (n = 32,041) with HPO Mapper Protocol 2, converting findings into HPO terms and linked genes. The cohort-level phenotype distribution reflects expected IBD biology, with Crohn’s disease and Ulcerative colitis among the most frequent terms, alongside Iron deficiency anaemia, Gastritis, Gastrointestinal ulcer, Drug allergy, Abdominal pain and Other gastrointestinal symptoms (Figure 4A). These results demonstrate that routine hospital text contains rich, recoverable phenotypic signals that can be normalised to HPO terms at scale.

**Figure 4:**
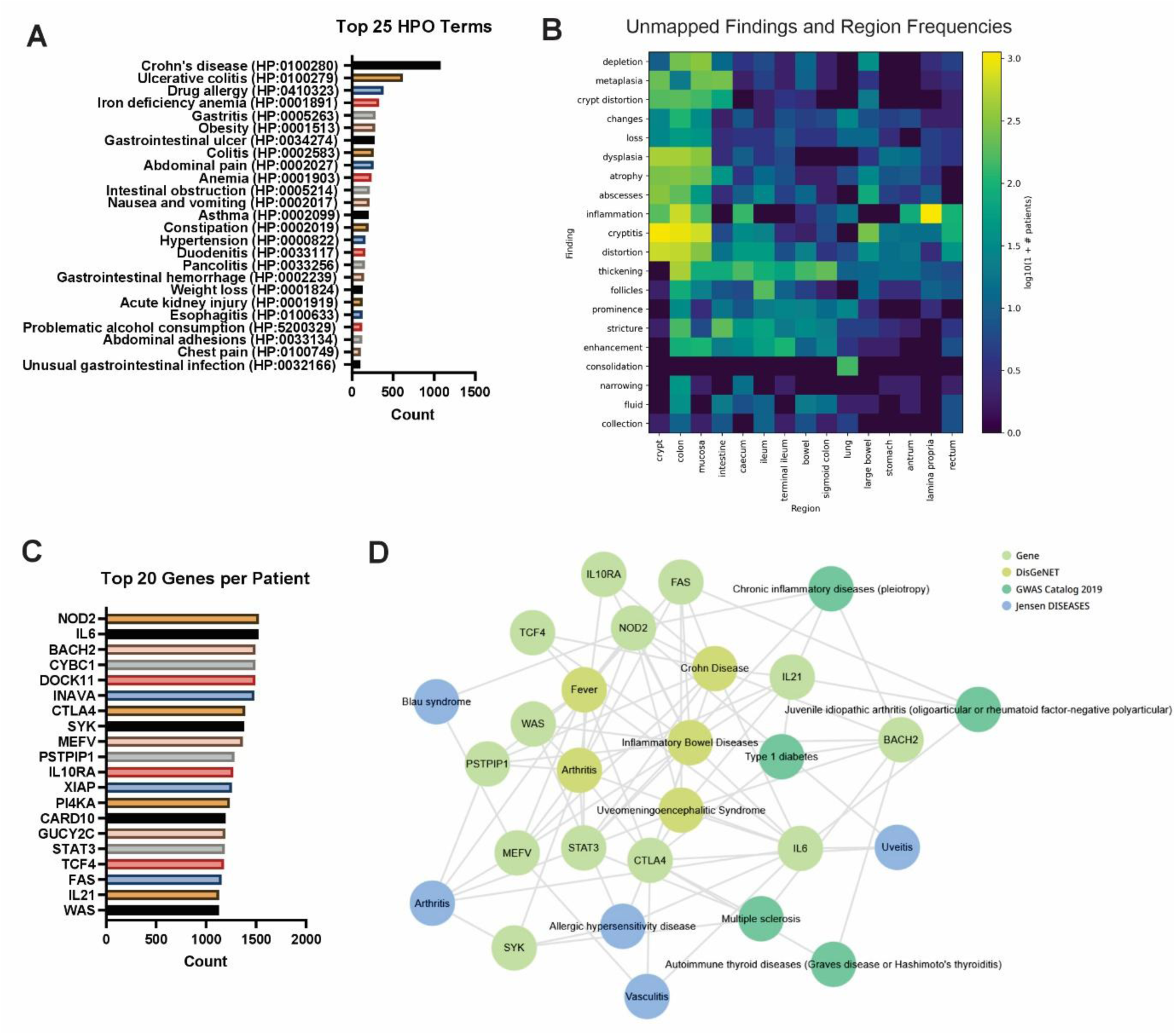
HPO Mapper Protocol 2 can map patient histology and radiology reports to HPO terms. A) Schematic of IBD cohort (n = 1,837) derived histology and radiology written reports (n = 32,041) structured using an IBD-adapted LLM into findings and regions to be mapped by HPO Mapper Protocol 2. B) Top 20 mapped and C) unmapped HPO terms using finding and regions from clinical reports using HPO Mapper Protocol 2. D) Enrich-KG knowledge graph of top 20 genes pulled from HPO terms for DisGeNET, GWAS Catalog 2019, and Jensen DISEASES.

Specific finding and region pairs that failed to map highlight concentrated gaps in HPO, most prominently observed for inflammation-related descriptors and histology-driven features such as cryptitis, thickening and dysplasia across colon, rectum, terminal ileum and related subregions (Figure 4B). These unmapped clusters suggest specific areas where ontology coverage or synonym handling could be strengthened, particularly for region-qualified histopathology phrases that are common in IBD reporting.

Associating HPO terms with their curated gene annotations and quantifying the most frequently implicated genes, the resulting ranked list included *NOD2, IL6, BACH2, CTLA4, STAT3, IL10RA, IL21, XIAP, MEFV* and *SYK* (Figure 4C). These top genes have high relevance to inflammatory pathways and related disease concepts, including inflammatory bowel disease, Crohn disease, chronic inflammatory disorders, arthritis, uveitis, multiple sclerosis, type 1 diabetes and Blau syndrome (Figure 4D). Together, these results show that HPO-normalised phenotypes can be projected onto gene and pathway space to recover known mechanisms in IBD and to generate testable hypotheses for patient stratification and target discovery.

### HPO Mapper extracted terms can improve causal gene list generation for patients

To demonstrate the utility of HPO Mapper extracted terms, we obtained histology and radiology reports from three Crohn’s disease patients with clinically validated mutations in *CTLA4*, *XIAP*, and *FOXP3*, obtained HPO terms, and calculated a patient-gene HPO overlap score (Figure 5A). Across each patient, we obtained and average of 53 HPO terms ranging from Type I diabetes mellitus, Sepsis, and Pancytopenia (Supplementary Table 1). Within each of the three clinically diagnosed patient, each confirmed causal gene was observed within the top 0.70% of all genes with HPO annotations (n = 5,252). Specifically, *CTLA4* ranked 21^st^ (Figure 5B), *XIAP* ranked 37^th^ (Figure 5C), and *FOXP3* ranked 7^th^ (Figure 5D).

**Figure 5:**
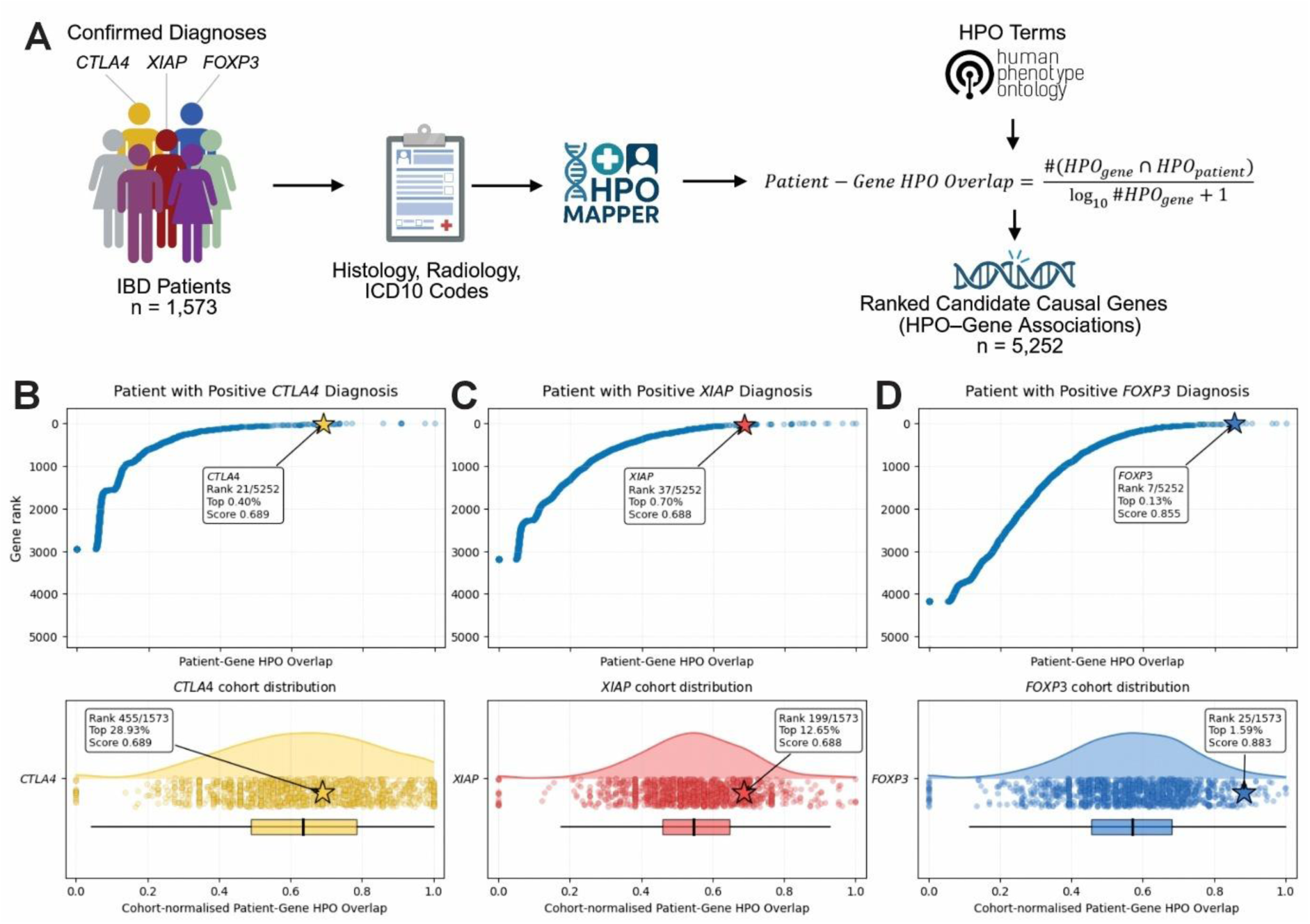
Mapped HPO terms allow improved detection of causal gene detection. A) Schematic of clinically diagnosed patients with mutations in *CTLA4*, *XIAP*, and *FOXP3* extracted HPO terms using HPO Mapper. B) Inflammatory bowel disease patients diagnosed with high HPO term overlap and positive diagnosis in B) *CTLA4*, C) *XIAP* and D) *FOXP3* showing gene rank vs patient-gene HPO overlap and respective patient-gene HPO overlap score distribution among cohort (n = 5,252). Box plots display median with IQR.

Among our entire cohort of IBD patients, both *CTLA4* and *XIAP-*diagnosed patients showed a modest cohort-normalised signal (top 28.93% and 12.65% respectively), with the *FOXP3*-diagnosed patient scoring within the top 1.59% of all patients, suggesting a more distinct phenotype. These finding demonstrated that phenotypic terms obtained from clinical records using HPO Mapper can be used to help prioritise causal genes in patients.

## DISCUSSION

Our results demonstrate embedding-based semantic mapping, coupled with LLM quality control, is a robust and scalable approach for clinical phenotyping. Unsupervised embeddings of the HPO preserved biological structure and clustered organ and system terms in a way that reflected known disease biology, a methodology which has been previously explored in similar use cases [31,37]. Like other studies, we validate cosine-similarity matching over an embedding space that carries meaningful semantics and provides a principled basis for downstream mapping rather than relying only on lexical overlap[37–39].

Head-to-head evaluation on a manually curated real-world IBD patient dataset confirmed that using LLMs for QC checking or to select top HPO terms improves mapping quality (Figure 1D). Recent studies have demonstrated that LLM-augmented phenotyping pipelines outperform rule-based pipelines for HPO extraction and normalisation, typically boosting precision while maintaining recall[40]. In our study, introducing an LLM QC step (Protocol 2) increased precision while maintaining recall, largely by filtering false positives that arise from near-neighbour but semantically inappropriate candidates. At cohort scale, HPO Mapper translated 50,489 ICD10 codes from 1,837 patients into HPO terms with higher coverage and a larger set of unique mappings than either a direct ICD10 to SNOMED CT to HPO mapping or BioPortal Annotator. Further, allowing the LLM to assist with selection (Protocol 3) delivered small additional gains, although Protocol 2 provided the best balance of precision and recall. Importantly, Protocol 2 (HPO Mapper with LLM QC) also yielded the greatest number of unique HPO terms, indicating not only improved recall and preserved precision but also an expanded phenotypic vocabulary. This increase in term diversity enhances biological granularity by capturing subtler and more specific phenotype distinctions that are missed by traditional mappings or automated annotators.

Compared to other contemporaneous tools, HPO Mapper P2 demonstrated adequate performance when starting with either ICD10 codes or extracted finding/region pairs (Table 4). Notably, HPO-RAG, Fenominal, and FastHPOCR can pull HPO terms directly from unstructured free-text[16,17,31], whereas HPO Mapper requires extracted finding/region pairs (as previously described[24]). Computational efficiency, which was comparable between HPO-RAG and HPO Mapper (Supplementary Figure 3), is critical for clinical and research settings where privacy constraints favour on-premises or secure deployment, and GPU access is limited. Our pipeline was designed to reduce computational load by precomputing HPO embeddings once and using a lightweight, fine-tuned LLM only for short QC prompts. In practice this reduces latency, infrastructure cost and carbon footprint, and makes large-scale runs feasible for tens of thousands of inputs[41–43]. Although larger models can provide marginal gains, the near-parity of smaller instruction-tuned models for QC argues for lightweight options in production. Further efficiency can be achieved with quantisation to 4–8-bit weights, batched inference, prompt caching, rule-based prefilters and knowledge-distilled variants of stronger models[44]. These choices are important for routine hospital deployments where throughput, cost control and data residency are as important as accuracy.

We estimate that a dual-A100 node consumes roughly 0.8 kWh per hour, equating to about 100 g of CO₂ emissions (data not shown). Therefore, a 60-hour job produces approximately 6 kg of CO₂. Although this is small compared with other clinical activities, for example, a single MRI scan emits ∼14.6 kg CO₂ and operating one MRI scanner for a year produces ∼58 tonnes CO₂, the cumulative impact becomes relevant when such workloads are executed repeatedly or scaled across institutions[45]. This underscores the value of lightweight models and efficient pipelines in reducing the environmental impact of clinical AI workloads.

Improvement in phenotypic mapping has the potential to increase the interpretation and identification of genetic disease[46,47]. We demonstrate the ability for HPO terms, derived from routinely collected clinical data to map to biologically relevant genes within IBD. This ability unlocks the potential to refine ontological terms and their link to specific genetic risk factors or causes. The overall benefit of accurately extracting these clinical data at the patient level for integration with the same patient’s genomics data is the ability to enable genuinely personalised diagnostics[48]. Such integration strengthens the resolution of phenotype-genotype relationships and supports the identification of single-gene monogenic primary immunodeficiencies masquerading as complex inflammatory disorders (e.g., IBD)[49–51].

By linking precise, patient-specific phenotypic signatures to their underlying genetic drivers, this approach improves diagnostic accuracy, accelerates recognition of rare monogenic disease, and directly informs more targeted clinical management[46]. In our study, HPO Mapper-derived HPO terms was able to generate clinically useful gene lists which would significantly improve identification in causal genes (Figure 5). We expect that, in an unbiased, non-disease-specific cohort, such as a population-scale resource with access to complete medical records[52], this approach could further improve identification of the true monogenic driver of disease. Rather than relying on disease-specific ascertainment, pathogenic variants in genes not traditionally annotated with IBD phenotypes would be prioritised more accurately. Conversely, for patients with relevant inflammatory phenotypes, monogenic IBD-associated genes such as *XIAP* and *CTLA4* would be expected to rank more highly across a non-disease specific cohort, strengthening unbiased gene-phenotype discovery and diagnostic interpretation.

Although we attempt to address annotation-density bias in the patient-gene HPO overlap score, improved phenotype to gene coverage within HPO would improve analyses. For example, well-studied genes such as *TP53*, *BRCA1/2*, and *CFTR* have far richer HPO annotation than less-characterised genes[1], potentially inflating their rank as candidate causal genes. Future versions could incorporate improved gene-level annotation normalisation and expand gene-phenotype mappings using resources such as OMIM, ClinGen, Orphanet, and literature-derived evidence. Patient phenotyping could also be strengthened by integrating longitudinal clinical records beyond imaging, histology, and ICD-10 data. Thus, HPO Mapper presents clear translational benefit, both in the identification of novel phenotype-genotype associations and in the recognition of rare monogenic disease masquerading as complex disease.

This study has several limitations. First, all primary evaluations used IBD data from a single health system, so generalisability to other specialties, institutions and languages remains to be demonstrated. Second, the manually curated benchmark (testing set) comprised 822 finding-region pairs, which is sufficient for protocol comparison but still modest for definitive ranking of closely performing models. Third, our ICD10 experiments inherit documentation and coding biases, and the direct ICD10 to SNOMED CT to HPO baseline is restricted by the coverage of available ontology mappings. Fourth, some clinically common histopathology phrases that combine finding and region were unmapped, indicating gaps in ontology coverage and synonym handling. Fifth, we did not fully model temporality, uncertainty, or nuanced negation in free text, which can influence phenotype assignment.

Addressing these limitations will involve multi-site validation, expansion of synonym and phrase libraries for region-qualified terms, multilingual evaluation, explicit handling of negation and time, and active-learning loops with clinical curators.

## CONCLUSIONS

HPO Mapper bridges structured and semi-structured clinical inputs to standard phenotype ontologies using modern AI while remaining practical for real-world deployment. Through pairing biologically meaningful embeddings with lightweight LLM mediation, it enables accurate, interpretable, and scalable phenotyping that supports genotype-phenotype investigation and precision medicine. By converting routine clinical records into consistent HPO profiles with improved coverage and reliability, HPO Mapper enables richer patient-level phenotyping that can be directly linked to genes and pathways for stratification. This creates a practical foundation for personalised medicine, supporting risk and subtype definition, genotype-guided prioritisation, and scalable cohort-to-individual translation across diverse clinical datasets.

## AVAILABILITY AND REQUIREMENTS

**Project name:** HPO Mapper

**Project home page:** https://huggingface.co/spaces/UoS-HGIG/HPOmapper;

https://github.com/UoS-HGIG/HPO-Mapper

**Operating system(s):** Platform independent

**Programming language:** Python

**License:** MIT

**Any restrictions to use by non-academics:** none

## Data Availability

Due to patient privacy restrictions, underlying clinical data cannot be publicly shared. Therefore, we are providing a user-friendly demo available at https://huggingface.co/spaces/UoS-HGIG/HPOmapper and https://github.com/UoS-HGIG/HPO-Mapper, containing underlying code and data. Any additional information required to reanalyse the data reported in this work paper is available from the lead contact upon request.

https://github.com/UoS-HGIG/HPO-Mapper

## LIST OF ABBREVIATIONS

AI: artificial intelligence
HPO: Human Phenotype Ontology
IBD: inflammatory bowel disease
IBDU: inflammatory bowel disease unclassified
ICD-10: International Statistical Classification of Diseases and Related Health Problems, 10th Revision
NHS: National Health Service
REC: Research Ethics Committee
GA4GH: Global Alliance for Genomics and Health
ISO: International Organization for Standardization
NLP: natural language processing
NCBO: National Center for Biomedical Ontology
LLM: large language model
QC: quality control
P1/P2/P3: Protocol 1/Protocol 2/Protocol 3
API: application programming interface
JSON: JavaScript Object Notation
PCA: principal component analysis
t-SNE: t-distributed stochastic neighbour embedding
L2: Euclidean (L2) normalization
SEM: standard error of the mean
FDR: false discovery rate
TP/FP/FN: true positive/false positive/false negative
SNOMED CT: Systematized Nomenclature of Medicine Clinical Terms
HGNC: HUGO Gene Nomenclature Committee
KEGG: Kyoto Encyclopedia of Genes and Genomes
DisGeNET: disease-gene association database
GWAS: genome-wide association study
MRI: magnetic resonance imaging
CT: computed tomography
CPU: central processing unit
GPU: graphics processing unit
HPC: high-performance computing
kWh: kilowatt-hour
CO₂: carbon dioxide
2D/3D: two-dimensional/three-dimensional gene symbols
NOD2: nucleotide-binding oligomerization domain containing 2
IL6: interleukin 6
STAT3: signal transducer and activator of transcription 3
IL10RA: interleukin 10 receptor subunit alpha
CTLA4: cytotoxic T-lymphocyte associated protein 4
BACH2: BTB domain and CNC homolog 2
IL21: interleukin 21
XIAP: X-linked inhibitor of apoptosis
MEFV: MEFV innate immunity regulator (pyrin)
SYK: spleen associated tyrosine kinase.

## DECLARATIONS

### Ethics approval and consent to participate

We obtained clinical data from a total of 1,837 research-consented patients diagnosed with inflammatory bowel disease recruited through the University of Southampton paediatric and adult clinics as part of the ethically approved Southampton Genetics of IBD Study (REC 09/H0504/125). All consenting participants meeting these criteria were included without further sampling or selection.

### Availability of data and materials

Due to patient privacy restrictions, underlying clinical data cannot be publicly shared. Therefore, we are providing a user-friendly demo available at https://huggingface.co/spaces/UoS-HGIG/HPOmapper and https://github.com/UoS-HGIG/HPO-Mapper, containing underlying code and data. Any additional information required to reanalyse the data reported in this work paper is available from the <u>lead contact</u> upon request.

### Authors’ contributions

AZK contributed to conceptualisation, formal analysis, investigation, methodology, software, validation, visualisation, writing the original draft, and writing (review and editing); ZG contributed to data curation, investigation, methodology, validation, and writing (review and editing); AB, MG, AH, MS, and RMB contributed to resources (with RMB also contributing to supervision); CMK contributed to supervision; PR provided supporting contributions to investigation and methodology and contributed equally to writing (review and editing); JJA contributed to conceptualisation, funding acquisition, investigation, methodology, resources, supervision, and writing (review and editing); and SE contributed to conceptualisation, formal analysis, funding acquisition, investigation, methodology, project administration, resources, supervision, and writing (review and editing). All authors read and approved the final manuscript.

## Acknowledgements

The authors acknowledge the IRIDIS High Performance Computing Facility at the University of Southampton, the Southampton Emerging Therapies and Technologies (SETT) Centre data and AI team, the Clinical Informatics Research Unit (CIRU), and the Southampton NIHR Biomedical Research Centre (BRC). Above all, we would like to thank the patients and their families.

