## Supplementary Figures for "Human Phenotype Ontology (HPO) Mapper: Semantic Mapping of Clinical Findings to the Human Phenotype Ontology Using AI-Powered Embeddings and LLM-Based Quality Control"

**A**

**ICD10 Code Nomenclature**

| | Category | | | | Etiology/Anatomy/Severity | | | Extension |
|---|---|---|---|---|---|---|---|---|
| | 1st | 2nd | 3rd | | 4th | 5th | 6th | 7th |
| Injury of intra-abdominal organs | S | 3 | 6 | . | | | | |
| Injury of liver | S | 3 | 6 | . | 1 | | | |
| Laceration of liver | S | 3 | 6 | . | 1 | 1 | | |
| Laceration of liver, unspecified degree | S | 3 | 6 | . | 1 | 1 | 3 | |
| Laceration of liver, unspecified degree, initial encounter | S | 3 | 6 | . | 1 | 1 | 3 | A |

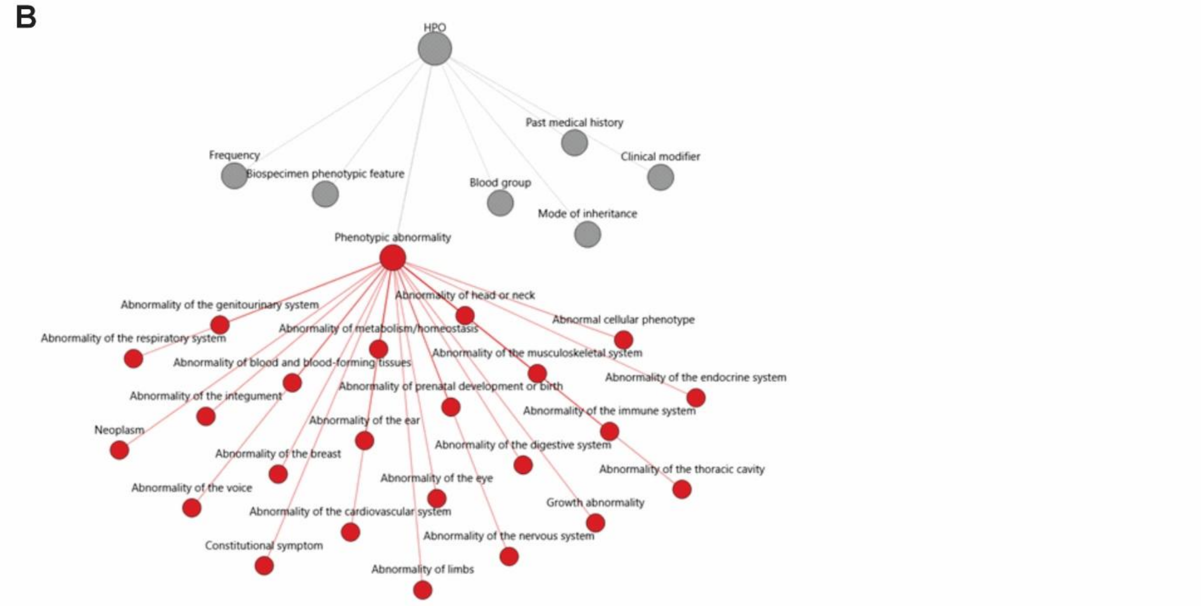


**Supplementary Figure 1: ICD10 and HPO overview.** A) Nomenclature of ICD10 codes including category, aetiology/anatomy/severity, and extension. B) Graphical representation of human phenotype ontology (HPO) top-level terms including Phenotypic abnormality branch used for visualisation.

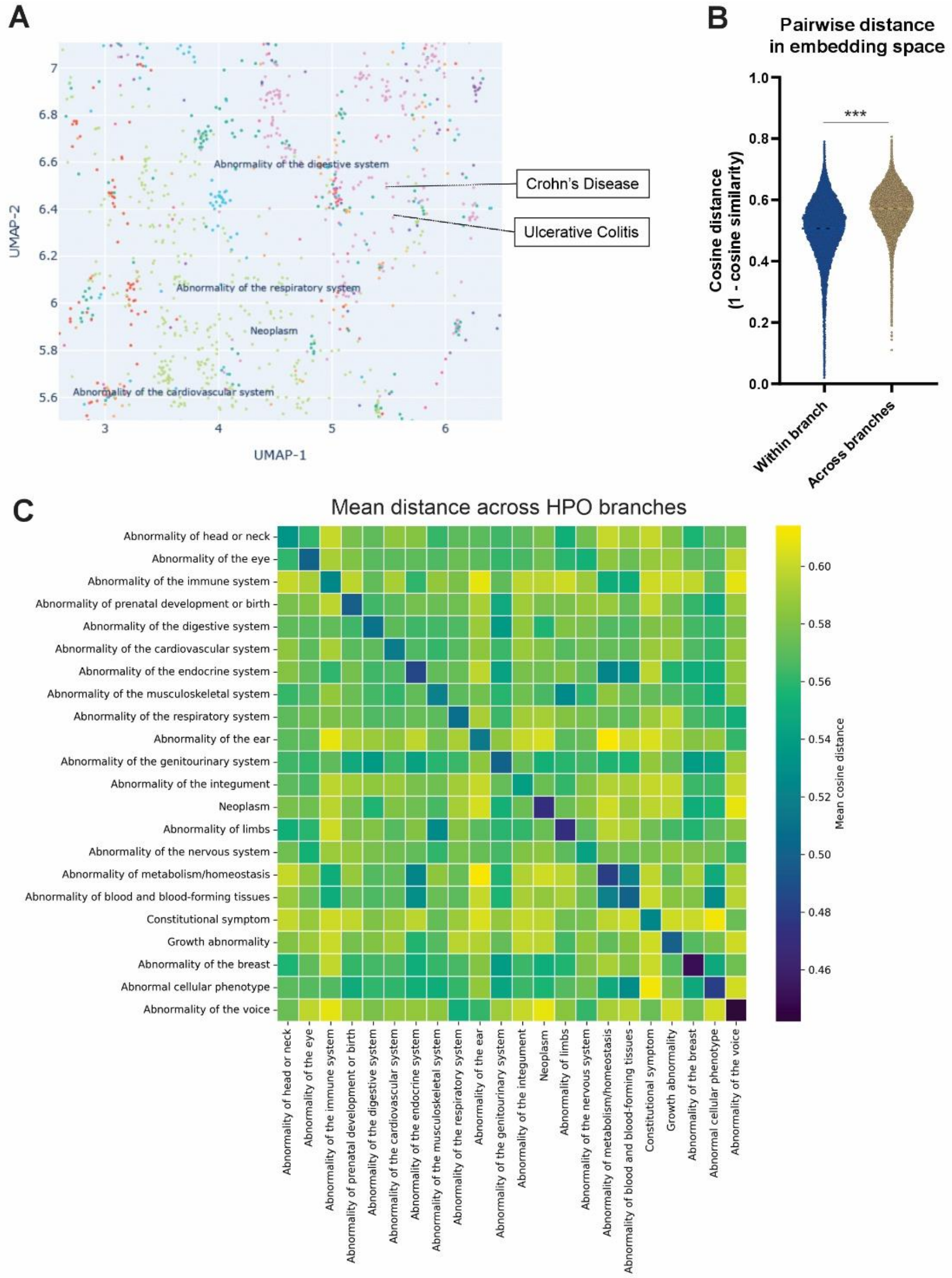


**Supplementary Figure 2: Semantic embeddings of HPO formed biologically relevant clusters.** A) Zoom of "Abnormality of the digestive system" (HP:0025031) showing location of "Crohn's Disease" and "Ulcerative Colitis" embeddings. B) Violin plots of cosine distance (1 − cosine similarity) computed in the original high-dimensional embedding. C) Heat map of mean cosine distance between top-level categories under HP:0000118 computed in the original embedding space.

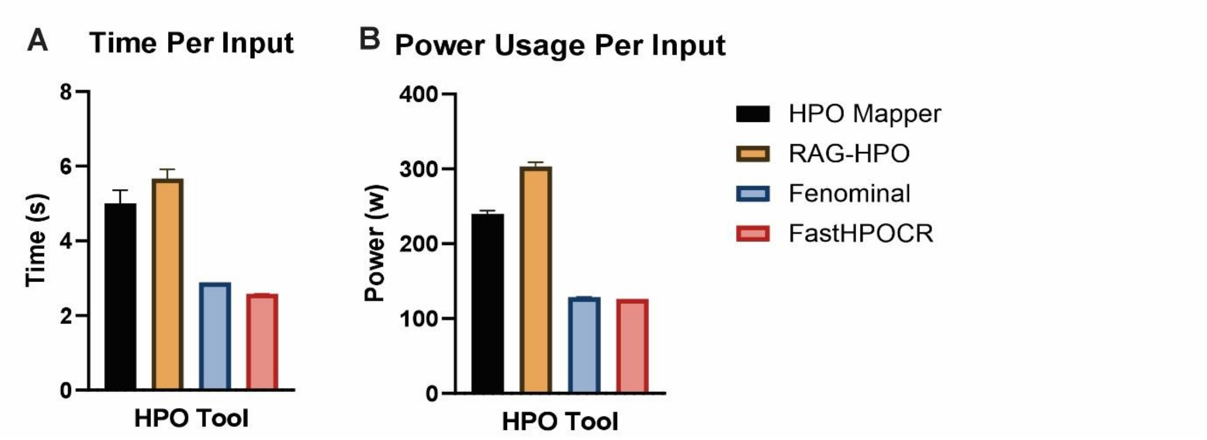


**Supplementary Figure 3: HPO Conversion Tool Time and Power Consumption Comparison.** Comparison of HPO Mapper, RAG-HPO, Fenominal, and FastHPOCR to process input text (ICD10 codes and finding/region pairs) as measured by A) average time in seconds and B) average power consumption in watts. All data are shown as mean ± SEM

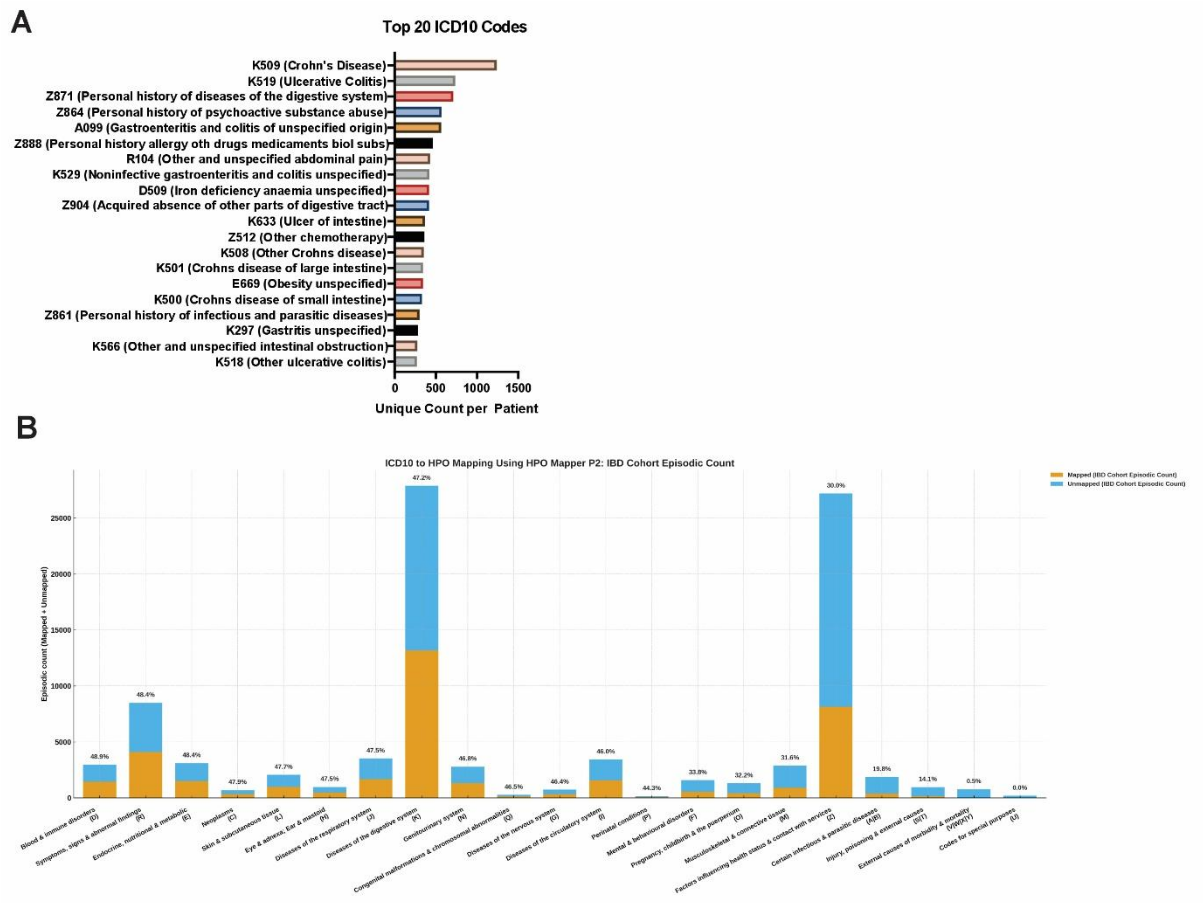


**Supplementary Figure 4: IBD Cohort ICD10 Code Conversion to HPO.** A) Top 20 ICD10 codes (unique per patient) obtained from patient clinical records. B) Mapping coverage of ICD10 to HPO conversion using HPO Mapper P2 by ICD10 prefix for all ICD10 codes in IBD cohort.
